# Study of Epidemiological Characteristics and In-silico Analysis of the Effect of Interventions in the SARS-CoV-2 Epidemic in India

**DOI:** 10.1101/2020.04.05.20053884

**Authors:** Archisman Mazumder, Vishwesh Bharadiya, Parul Berry, Mehak Arora, Mudit Agarwal, Mohak Gupta, Giridara Gopal Parameswaran, Priyamadhaba Behera

## Abstract

After SARS-CoV-2 set foot in India, the Government took a number of steps to limit the spread of the disease in the country. This study involves assessing how the disease affected the population in the initial days of the epidemic. Data was collected from government-controlled and crowdsourced websites and analyzed. Studying age and sex parameters of 413 Indian COVID-19 patients, the median age of the affected individuals was found to be 36 years (IQR, 25-54) with 20-39 years males being the most affected group. The number of affected males (66.34%) was more than that of the females (33.66%). Using Susceptible-Infected-Removed (SIR) model, the range of contact rate (β) of India was calculated and the role of public health interventions was assessed. If current contact rate continues, India may have 5583 to 13785 active cases at the end of 21 days lockdown.

**Article Summary Line:** The study gives the epidemiological characteristics of the SARS-CoV-2 epidemic in India, where unlike other countries, the 20-39 years males are most affected, and the SIR model predicts the probable number of cases of COVID-19 by the end of the 21 days lockdown in the country, which will help to develop appropriate public health interventions to control the COVID-19 epidemic.

## INTRODUCTION

In December 2019, a cluster of patients with pneumonia of unknown origin was encountered in Wuhan, China *(1)*. The causative agent was determined to be a novel virus of the *Coronaviridae* family of RNA viruses and was claimed to be of zoonotic origin *(1)*. Due to its close relationship with the SARS-CoV, this novel coronavirus was named SARS-CoV-2 *(2)*. Since then, SARS-CoV-2 has caused a widespread outbreak of the disease now known as COVID-19 and was declared to be a pandemic by WHO on March 11 2020 *(3)*. Human to human transmission occurs primarily through close-contact with the infected person, through fomites in the immediate surroundings of the infected person and via droplets of respiratory secretions *(4, 5)*, although there is limited evidence pointing to a possibility of airborne and faeco-oral transmission as well *(6, 7)*. According to few case studies, transmission may also occur via viral shedding in “pre-symptomatic” individuals during the incubation period *(8, 9)*.

The incubation period for COVID-19 is thought to be within 14 days of exposure, with a median incubation period of 4-5 days *(4, 10, 11)*. The median age of patients affected by COVID-19 is 47 years with the most common clinical findings being fever and cough *(4, 12)*. Other symptoms include expectoration, headache, myalgia fatigue, and diarrhoea and haemoptysis in rare cases. About 18% of patients develop shortness of breath *(4)*. Severe disease has been reported to occur in 14% of patients with older age and pre-existing chronic disease being risk factors *(4, 13)*.

Critical disease requiring intensive care unit admission has been reported in 5 percent, and overall case-fatality rate as 2.3% *(13)*. Currently, there are no approved treatments for COVID-19 and clinical trials such as the WHO SOLIDARITY trial are underway to evaluate the effectiveness of drugs like lopinavir-ritonavir, remdesivir, hydroxychloroquine and azithromycin *(3, 14)*.

India reported its first case of COVID-19 on 30 January, 2020; a medical student who had travelled from Wuhan, China, the then epicenter of COVID-19 *(15)*. While there were only a few reported imported cases in the month of February, the number began to increase rapidly in March *(14)*. On 15th March 2020, India sealed its borders and stopped all international flights, meaning all the initial imported cases which seeded COVID-19 in India arrived in the Indian subcontinent before 15 March 2020 *(16)*. According to the data available in the public domain, India had approximately 3726 cases (both imported cases and due to person-to-person transmission) as of 5 April 2020 *(17)*. According to the Indian Council of Medical Research, as of 31st March 2020, community transmission has not yet started and India is in category 2 of WHO classification for transmission patterns, i.e., sporadic cases without evidence of community transmission *(18)*.

The most important question in the current scenario concerns the mathematical parameters of the initial spread of COVID-19 in India, and what are the epidemiological aspects that can predict this spread. We acknowledge there are certain difficulties in making precise calculations due to the rapidly changing dynamic of the epidemic in the early stages, limited availability of data in the public domain, absence of robust line listing of cases and limited testing capacity.

Nonetheless, mathematical models with reasonable assumptions based on available information can help in analysis of the currently available data to provide important insights for guiding public health interventions. For a predictive model to be relevant to the reality, the model itself should represent what is happening in the real world. The most basic of these models is the Susceptible-Infected-Removed (SIR) model *(19,20)*, which we’ve used in the current Indian scenario to determine the range in which contact rate β lies and also calculate the range of the current reproduction number, R_t_.

In the first part of our paper, we outline the several public health measures taken by India in response to the COVID-19 outbreak. We then study and analyze the epidemiological parameters of COVID-19 in India using publicly available data till 28 March 2020. Finally, we use the SIR model, and run simulations to determine the effective contact rate β, and derive from that the reproduction number R_t_, and comment on the result of the nationwide public health measures implemented from mid-March 2020 and the lockdown implemented from 25 March 2020.

## INDIA’S OUTBREAK RESPONSE

Since the beginning of the outbreak in India, there have been a number of interventions done at various levels by the state governments and by the Central Government agencies. The number of cases started increasing in March *(21)*, prompting a number of interventions to control the outbreak. The first interventions were mostly related to travel advisories and bans. Gradually there were more social distancing measures in March which were then followed by lockdowns, ultimately culminating in a nation-wide 21 days lockdown from 25 March, 2020 (Table 1)

**Table 1^a^:**
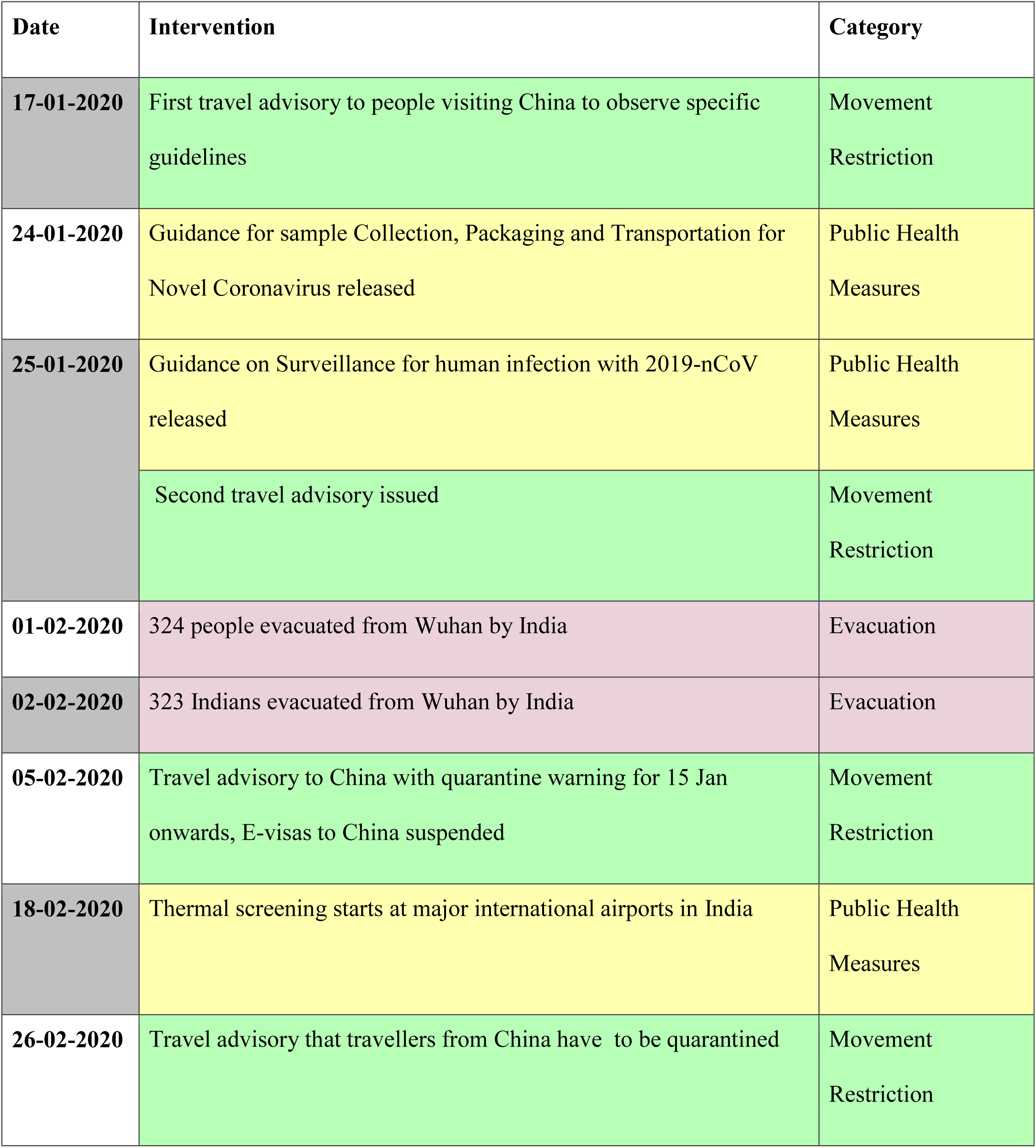
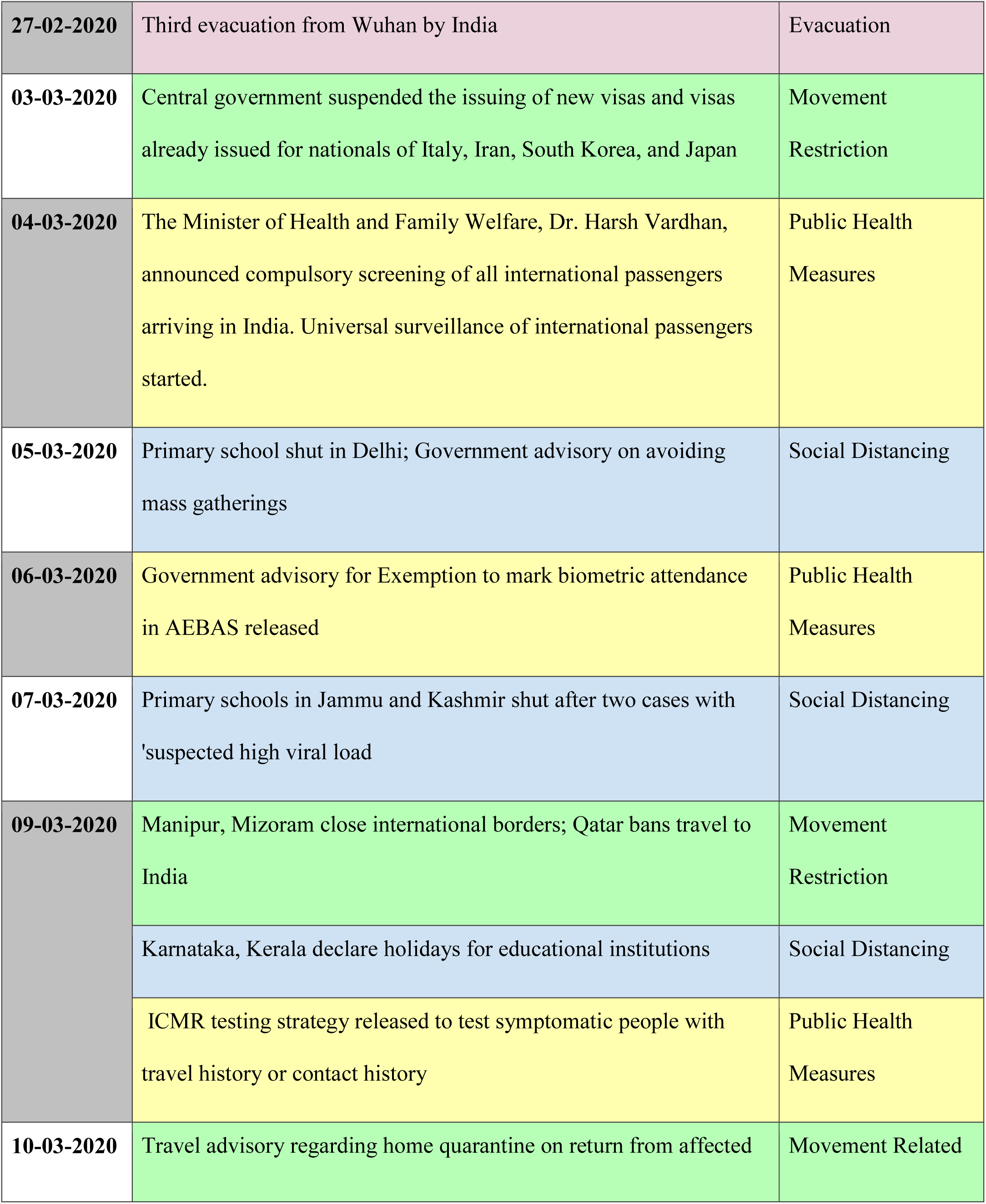
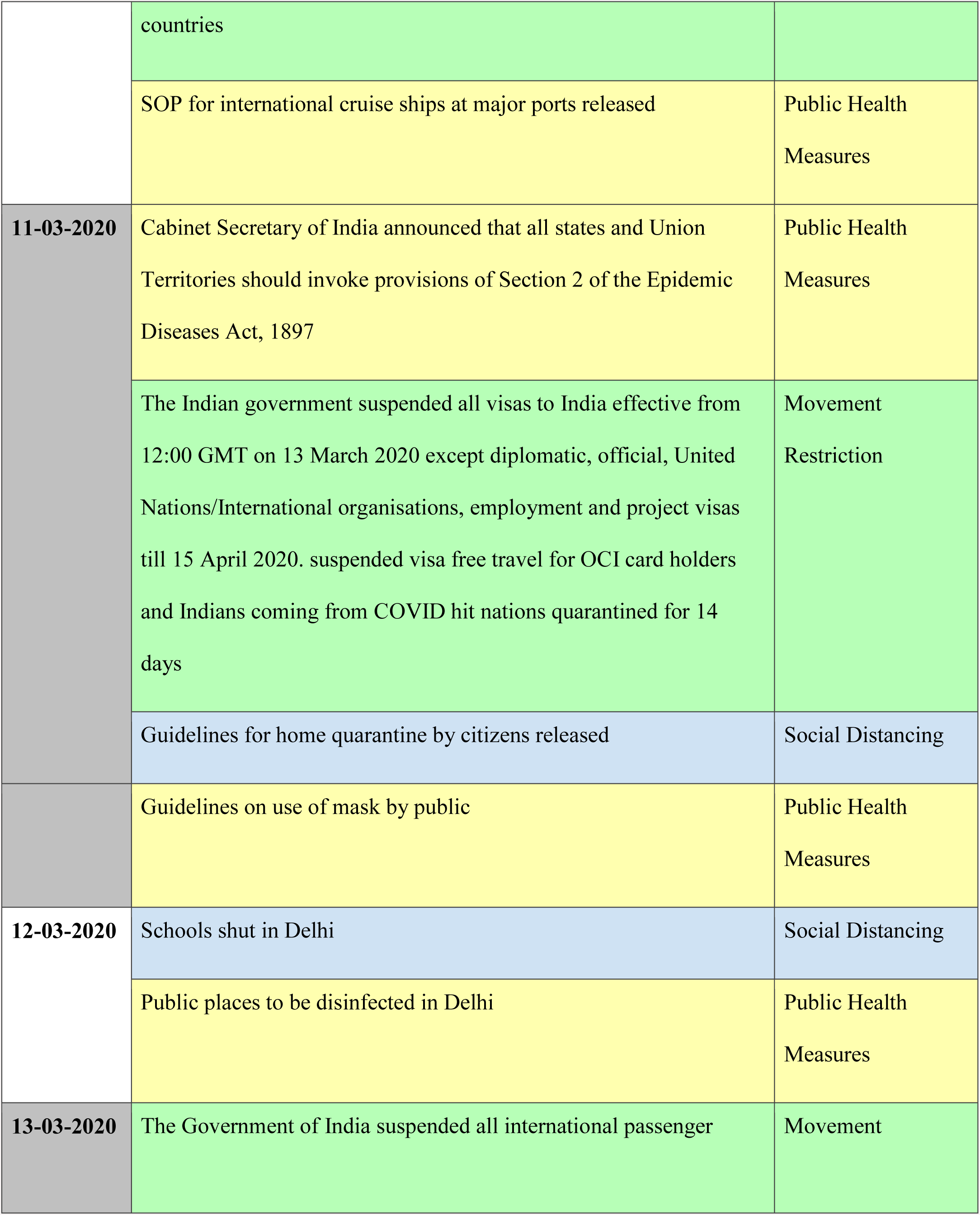
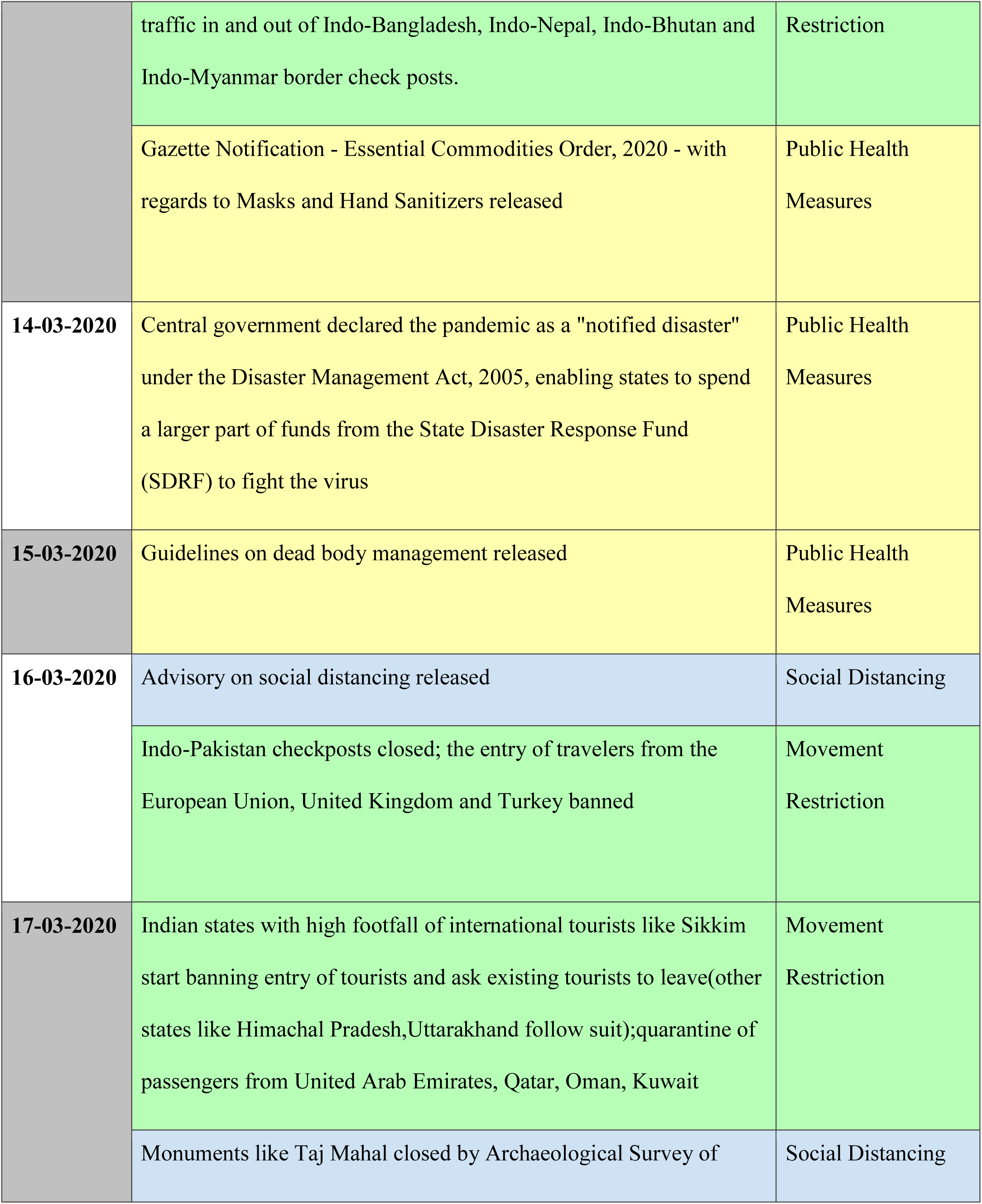
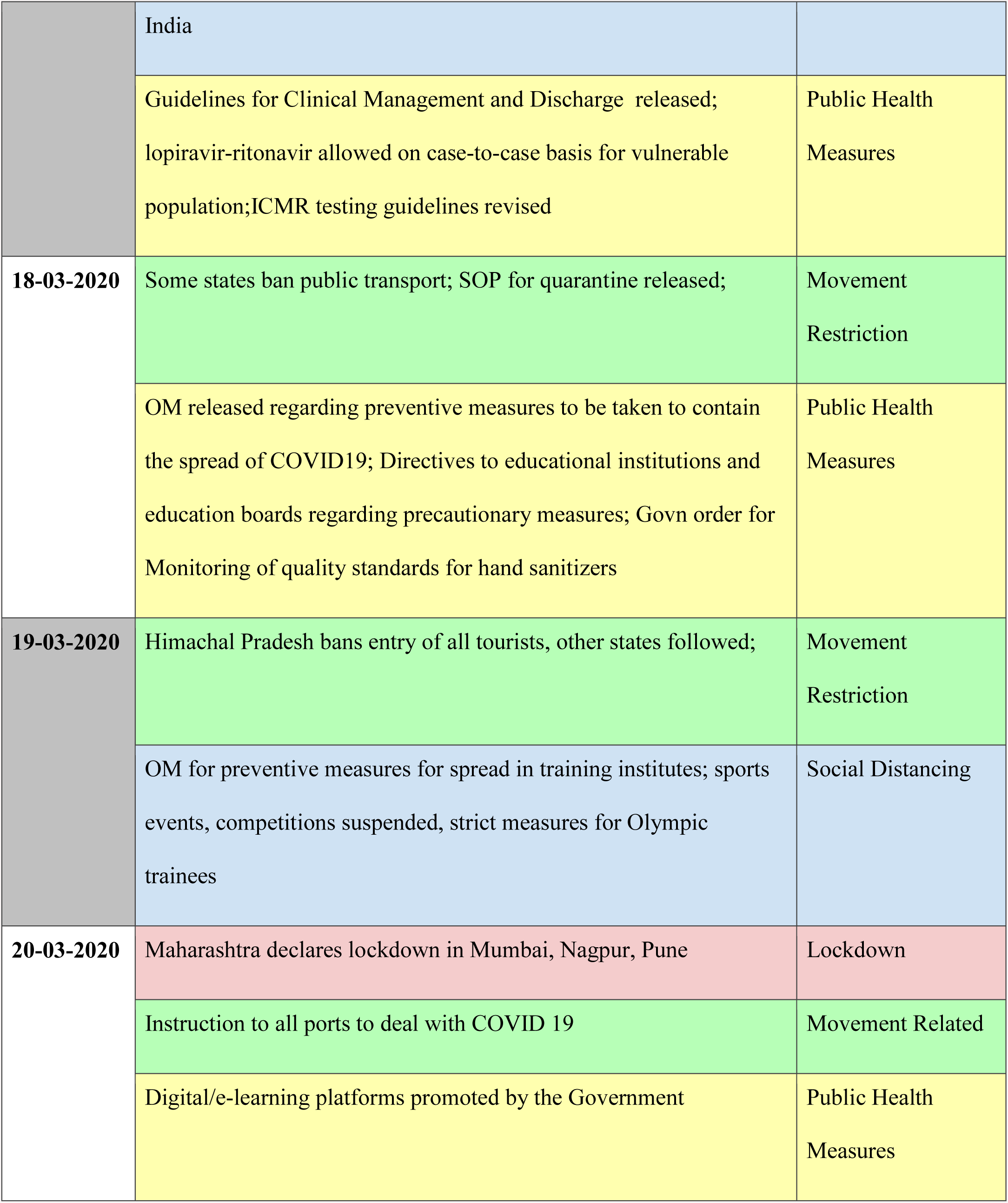
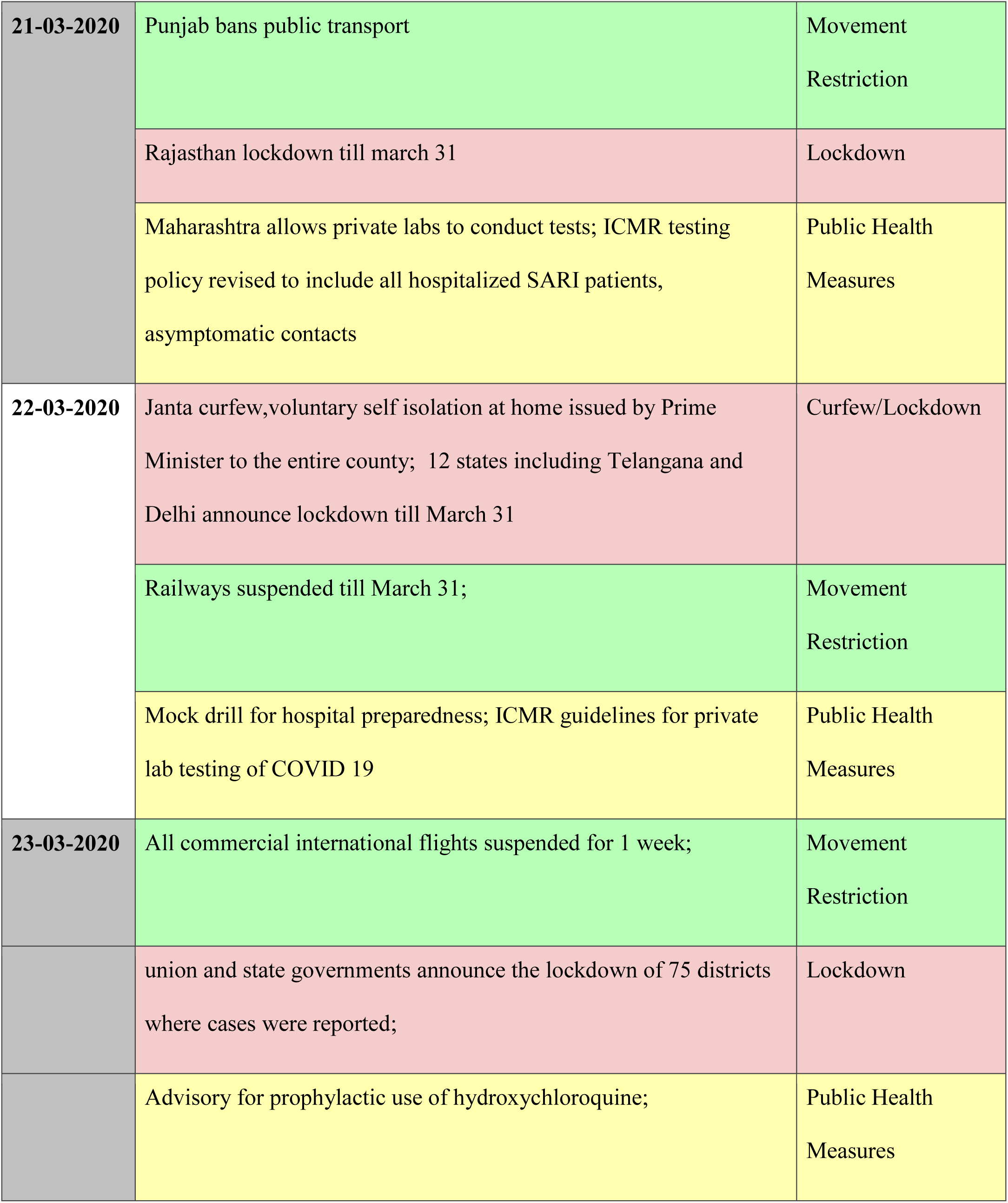
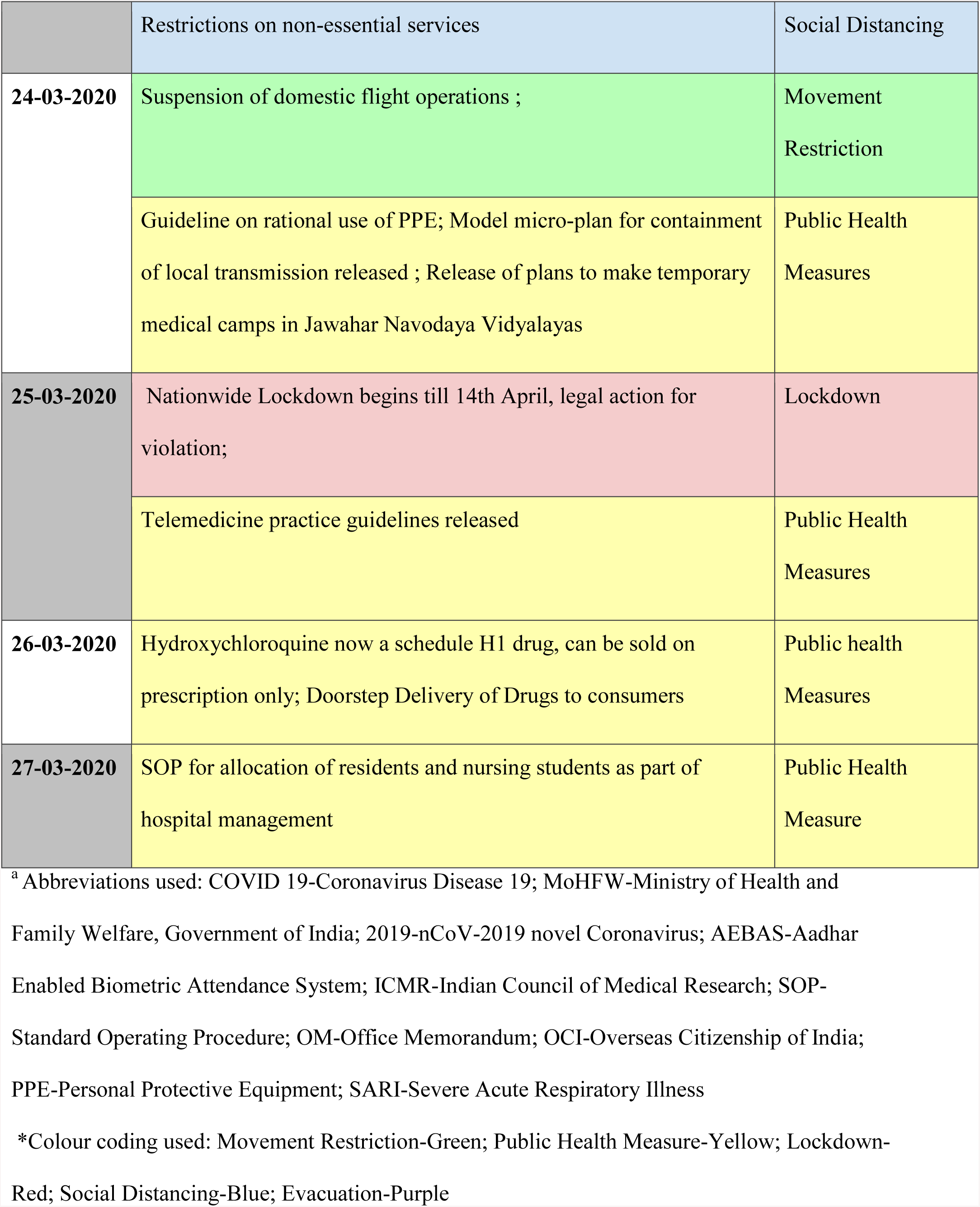
Interventions done at the central government or state level in India to control the COVID-19 outbreak till 27 March, 2020 (Source : MoHFW)*

| Date | Intervention | Category |
| --- | --- | --- |
| 17-01-2020 | First travel advisory to people visiting China to observe specific guidelines | Movement<br>Restriction |
| 24-01-2020 | Guidance for sample Collection, Packaging and Transportation for Novel Coronavirus released | Public Health<br>Measures |
| 25-01-2020 | Guidance on Surveillance for human infection with 2019-nCoV released | Public Health<br>Measures |
|  | Second travel advisory issued | Movement<br>Restriction |
| 01-02-2020 | 324 people evacuated from Wuhan by India | Evacuation |
| 02-02-2020 | 323 Indians evacuated from Wuhan by India | Evacuation |
| 05-02-2020 | Travel advisory to China with quarantine warning for 15 Jan onwards, E-visas to China suspended | Movement<br>Restriction |
| 18-02-2020 | Thermal screening starts at major international airports in India | Public Health<br>Measures |
| 26-02-2020 | Travel advisory that travellers from China have to be quarantined | Movement<br>Restriction |
| **27-02-2020** | Third evacuation from Wuhan by India | Evacuation |
| **03-03-2020** | Central government suspended the issuing of new visas and visas already issued for nationals of Italy, Iran, South Korea, and Japan | Movement<br>Restriction |
| **04-03-2020** | The Minister of Health and Family Welfare, Dr. Harsh Vardhan, announced compulsory screening of all international passengers arriving in India. Universal surveillance of international passengers started. | Public Health<br>Measures |
| **05-03-2020** | Primary school shut in Delhi; Government advisory on avoiding mass gatherings | Social Distancing |
| **06-03-2020** | Government advisory for Exemption to mark biometric attendance in AEBAS released | Public Health<br>Measures |
| **07-03-2020** | Primary schools in Jammu and Kashmir shut after two cases with 'suspected high viral load | Social Distancing |
| **09-03-2020** | Manipur, Mizoram close international borders; Qatar bans travel to India | Movement<br>Restriction |
|  | Karnataka, Kerala declare holidays for educational institutions | Social Distancing |
|  | ICMR testing strategy released to test symptomatic people with travel history or contact history | Public Health<br>Measures |
| **10-03-2020** | Travel advisory regarding home quarantine on return from affected | Movement Related |
|  | countries |  |
|  | SOP for international cruise ships at major ports released | Public Health Measures |
| **11-03-2020** | Cabinet Secretary of India announced that all states and Union Territories should invoke provisions of Section 2 of the Epidemic Diseases Act, 1897 | Public Health Measures |
|  | The Indian government suspended all visas to India effective from 12:00 GMT on 13 March 2020 except diplomatic, official, United Nations/International organisations, employment and project visas till 15 April 2020. suspended visa free travel for OCI card holders and Indians coming from COVID hit nations quarantined for 14 days | Movement Restriction |
|  | Guidelines for home quarantine by citizens released | Social Distancing |
|  | Guidelines on use of mask by public | Public Health Measures |
| **12-03-2020** | Schools shut in Delhi | Social Distancing |
|  | Public places to be disinfected in Delhi | Public Health Measures |
| **13-03-2020** | The Government of India suspended all international passenger | Movement |
|  | traffic in and out of Indo-Bangladesh, Indo-Nepal, Indo-Bhutan and Indo-Myanmar border check posts. | Restriction |
|  | Gazette Notification - Essential Commodities Order, 2020 - with regards to Masks and Hand Sanitizers released | Public Health Measures |
| **14-03-2020** | Central government declared the pandemic as a "notified disaster" under the Disaster Management Act, 2005, enabling states to spend a larger part of funds from the State Disaster Response Fund (SDRF) to fight the virus | Public Health Measures |
| **15-03-2020** | Guidelines on dead body management released | Public Health Measures |
| **16-03-2020** | Advisory on social distancing released | Social Distancing |
|  | Indo-Pakistan checkpoints closed; the entry of travelers from the European Union, United Kingdom and Turkey banned | Movement Restriction |
| **17-03-2020** | Indian states with high footfall of international tourists like Sikkim start banning entry of tourists and ask existing tourists to leave (other states like Himachal Pradesh, Uttarakhand follow suit); quarantine of passengers from United Arab Emirates, Qatar, Oman, Kuwait | Movement Restriction |
|  | Monuments like Taj Mahal closed by Archaeological Survey of | Social Distancing |
|  | India |  |
|  | Guidelines for Clinical Management and Discharge released; lopiravir-ritonavir allowed on case-to-case basis for vulnerable population;ICMR testing guidelines revised | Public Health Measures |
| 18-03-2020 | Some states ban public transport; SOP for quarantine released; | Movement Restriction |
|  | OM released regarding preventive measures to be taken to contain the spread of COVID19; Directives to educational institutions and education boards regarding precautionary measures; Govn order for Monitoring of quality standards for hand sanitizers | Public Health Measures |
| 19-03-2020 | Himachal Pradesh bans entry of all tourists, other states followed; | Movement Restriction |
|  | OM for preventive measures for spread in training institutes; sports events, competitions suspended, strict measures for Olympic trainees | Social Distancing |
| 20-03-2020 | Maharashtra declares lockdown in Mumbai, Nagpur, Pune | Lockdown |
|  | Instruction to all ports to deal with COVID 19 | Movement Related |
|  | Digital/e-learning platforms promoted by the Government | Public Health Measures |
| 21-03-2020 | Punjab bans public transport | Movement<br>Restriction |
|  | Rajasthan lockdown till march 31 | Lockdown |
|  | Maharashtra allows private labs to conduct tests; ICMR testing policy revised to include all hospitalized SARI patients, asymptomatic contacts | Public Health<br>Measures |
| 22-03-2020 | Janta curfew,voluntary self isolation at home issued by Prime Minister to the entire county; 12 states including Telangana and Delhi announce lockdown till March 31 | Curfew/Lockdown |
|  | Railways suspended till March 31; | Movement<br>Restriction |
|  | Mock drill for hospital preparedness; ICMR guidelines for private lab testing of COVID 19 | Public Health<br>Measures |
| 23-03-2020 | All commercial international flights suspended for 1 week; | Movement<br>Restriction |
|  | union and state governments announce the lockdown of 75 districts where cases were reported; | Lockdown |
|  | Advisory for prophylactic use of hydroxychloroquine; | Public Health<br>Measures |
|  | Restrictions on non-essential services | Social Distancing |
| 24-03-2020 | Suspension of domestic flight operations ; | Movement<br>Restriction |
|  | Guideline on rational use of PPE; Model micro-plan for containment of local transmission released ; Release of plans to make temporary medical camps in Jawahar Navodaya Vidyalayas | Public Health<br>Measures |
| 25-03-2020 | Nationwide Lockdown begins till 14th April, legal action for violation; | Lockdown |
|  | Telemedicine practice guidelines released | Public Health<br>Measures |
| 26-03-2020 | Hydroxychloroquine now a schedule H1 drug, can be sold on prescription only; Doorstep Delivery of Drugs to consumers | Public health<br>Measures |
| 27-03-2020 | SOP for allocation of residents and nursing students as part of hospital management | Public Health<br>Measure |

<sup>a</sup> Abbreviations used: COVID 19-Coronavirus Disease 19; MoHFW-Ministry of Health and
Family Welfare, Government of India; 2019-nCoV-2019 novel Coronavirus; AEBAS-Aadhar
Enabled Biometric Attendance System; ICMR-Indian Council of Medical Research; SOP-
Standard Operating Procedure; OM-Office Memorandum; OCI-Overseas Citizenship of India;
PPE-Personal Protective Equipment; SARI-Severe Acute Respiratory Illness
\*Colour coding used: Movement Restriction-Green; Public Health Measure-Yellow; Lockdown-
Red; Social Distancing-Blue; Evacuation-Purple

## METHODS

### Epidemiological analysis

#### Data Source

The raw data was collected from the patient database of covid19india.org which is a crowdsourced patient database for positive cases of SARS-CoV-2 which have been confirmed by laboratory testing, including data from state government and central government agencies (https://www.covid19india.org/). The data was taken for cases confirmed on 28 March, 2020 or earlier. Only those patient data were analyzed which had both age and sex data of the patient mentioned. For convenience of analysis, patients whose age data was given as a range were excluded.

#### Study population

The above method yielded a sample of size n=413. Of these patients, the status of the patient as of 28th March 2020 was not known for 13 patients.

#### Study Design

For the epidemiological analysis part of the study, we used a descriptive design. After collecting the data analysis was done in regard to the age distribution, status of patients and sex distribution using Microsoft Office Excel 2007. Fatality ratio in any category was found out by dividing the number of deaths in the category by the number of affected individuals of that category. Central tendencies for age of patients of various categories were also calculated.

### Mathematical analysis

The spread of the disease in the population is studied by the SIR model *(19,20)* which divides the (fixed) population of N individuals into three “compartments” which vary as a function of time (For purposes of this study, we have not included vital dynamics like birth and death rate.) –

- S(t) - S(t) are those susceptible but not yet infected with the disease (in a novel disease like nCOV-19,the entire population is assumed to be susceptible as there is no pre-existing immunity);
- I(t) - I(t) is the number of infectious individuals;
- R(t) - R(t) are those individuals who have been removed from the infected population(includes those who have recovered from the disease and also the deaths)

The SIR model describes the change in the population of each of these compartments in terms of two parameters, β and γ.

β-β describes the effective *contact rate* of the disease: a susceptible individual comes into contact with an infectious individual and acquires the disease. This parameter takes into account both the number of people contacted per unit time, and the effectiveness of transmission in each contact. It reflects the force of infection of the disease and helps us understand at what rate the epidemic is progressing

γ - γ is the mean *removal rate*: In our model, it is calculated using the removed cases as against the new cases on a daily basis.

β and γ are useful in the SIR model using the following differential equations-

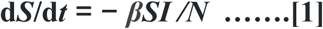

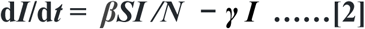

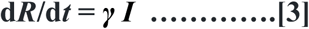

Assuming the present scenario in India where I<<N and R<<N, S is almost equal to N. So S/N=1. Putting this in equation 2 and integrating, we get

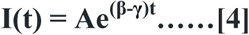

where A is determined the number of cases at t=0.

We have used the data available till 4th April to estimate the two parameters, β and R_t_ (time varying reproductive number) with the help of SIR model (www.statista.com). We assumed that the recovery rate **γ** would remain constant for the population. The removal rate followed a normal distribution, and the mean was calculated with the data available from 01 March to April 04, 2020 *(22)*. Since the effect of interventions would reflect in the contact rate β, we then took the value of **γ** to be constant equal to the mean (0.103) and ran the SIR model multiple times by varying the value of β, and comparing the trends with the real data. We plotted the trendline for the real data using Microsoft Office Excel 2007, and used the equation of the curve to find out the trend of β in India in the present day scenario by comparing it to equation 4.

## RESULTS

Among the 413 patients, there were 274 (66.34%) male and 319 (33.66%) female. The patients affected in India had a median age of 36 years (IQR, 25-54). The median age of females was higher (40 years; IQR, 24.5-59) than males (35 years; IQR, 25-50). Nearly half of the patients (49.88%) of affected patients were in the age group of 20-39 years. (refer Table 2). There were sixteen deaths (3.8%). Majority of them (68.75%) were in the 60-79 years age group. The median age of deceased patients was 65 years (IQR, 59.25-69). The mortality rate in male patients was 4.38% and for female patients was 2.88%. Overall case fatality rate was 3.87%. There is a 16.18% mortality rate in the 60-79 years age group and 20.0% in the equal to or above 80 years age group. (refer Table 3)

**Table 2:** Distribution of COVID-19 patients in India confirmed till 28th March 2020 across different age groups and sex (n=413)

| Age group | Male | Percentage of patients accounted (normalized value)* | Female | Percentage of patients accounted (normalized value)* |
| --- | --- | --- | --- | --- |
| <20years | 14 | 3.39%(0.18) | 12 | 2.91%(0.17) |
| 20-39 years | 149 | 36.08%(2.07) | 57 | 13.80%(0.88) |
| 40-59 years | 71 | 17.19%(1.53) | 37 | 8.96%(0.85) |
| 60-79 years | 38 | 9.20%(2.04) | 30 | 7.26%(1.54) |
| >=80 years | 2 | 0.48%(1.20) | 3 | 0.73%(1.82) |
| **Total** | **274** |  | **139** |  |

\*Normalized values have been calculated by dividing the percentage of patients in each category by the percentage of Indian population in that category.

**Table 3:** Distribution of COVID 19 patients of India with age, sex and status of disease updated till 28 March, 2020 (n=413)

| Variables | Deceased | Recovered | Hospitalized* | Status not known |
| --- | --- | --- | --- | --- |
| **AGE** |  |  |  |  |
| <20years | 0 | 2 | 22 | 2 |
| **20-39 years** | 2 | 5 | 195 | 4 |
| **40-59 years** | 2 | 4 | 98 | 4 |
| **60-79 years** | 11 | 2 | 52 | 3 |
| **>=80 years** | 1 | 1 | 3 | 0 |
| **GENDER** |  |  |  |  |
| **Male** | 12 | 11 | 243 |  |
| **Female** | 4 | 3 | 127 |  |
| **TOTAL** | **16** | **14** | **370** | **13** |

\*Hospitalized include patients in facility isolation, in ICU or under medical care in hospital

For recovered patients, most were in the 20-39 years age group followed by the 40-59 years age group. Median age was 36 years (IQR, 21.75-56.25). As per the data as on 28 March, 2020, 14 of these 413 patients had recovered.

The number of hospitalized patients accounted for 370 of the 413 patients. However, status of 13 of the 413 patients was not available.

After running multiple simulations using the SIR model, all assuming different values of β (Appendix Table), we found that the value of β in the current Indian scenario calculated from the trendline of real data lies around 0.258, which is also visible in the graph of real-time active cases lying between β=0.24 and β=0.29 curves (Figure 3).

**Figure 1.**
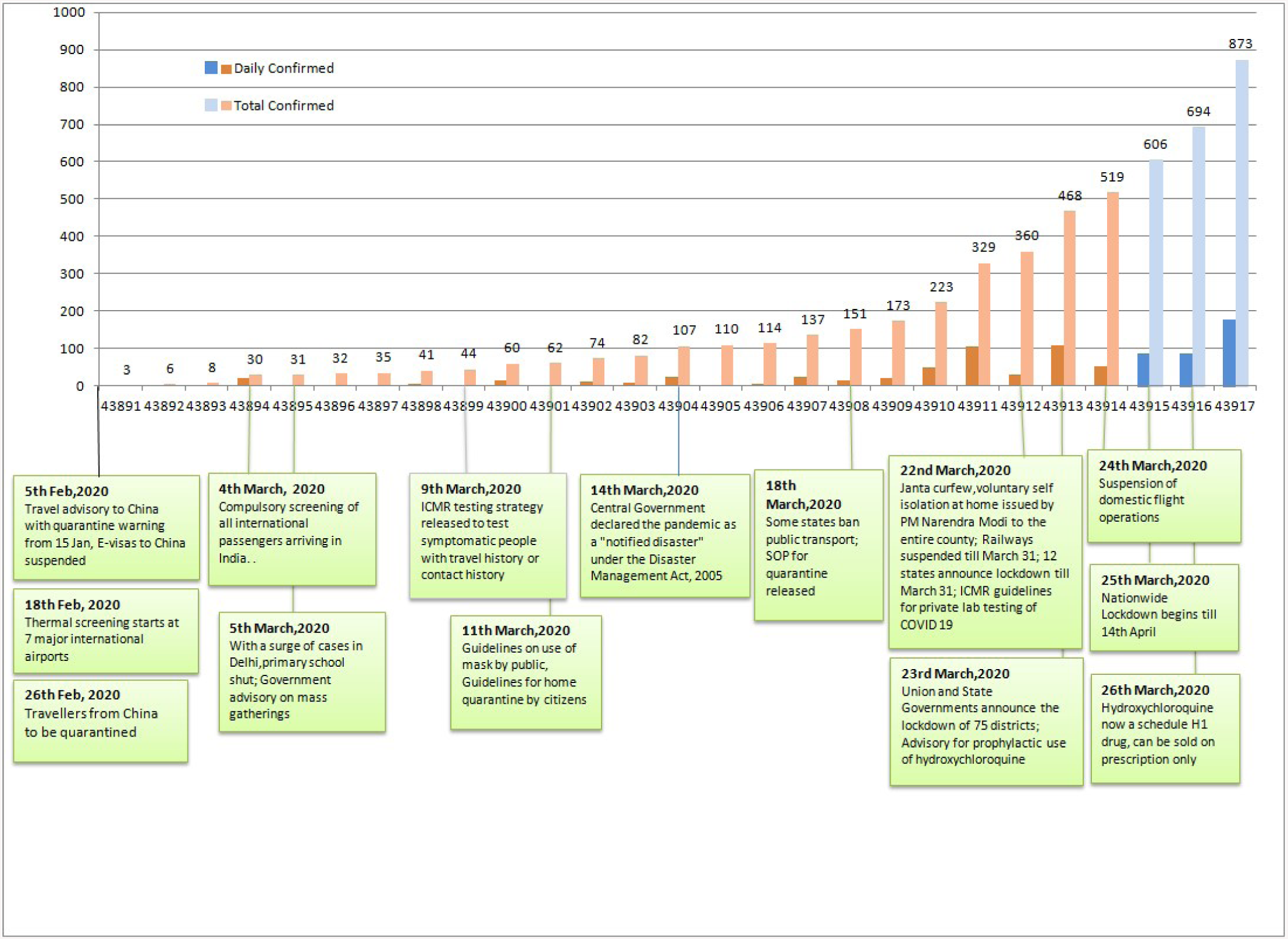
Day wise COVID-19 (new and cumulative) cases during 1^st^-27^th^ March, 2020 and the major interventions taken by the government to control the spread of the CoVID-19. Data Source: ICMR-NIE and MoHFW.

**Figure 2.**
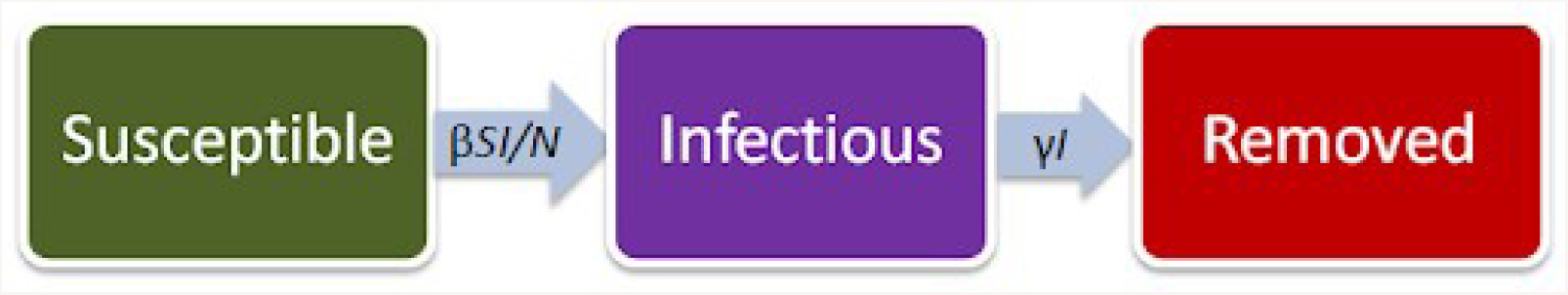
Flow diagram of transitions in SIR Model.

**Figure 3.**
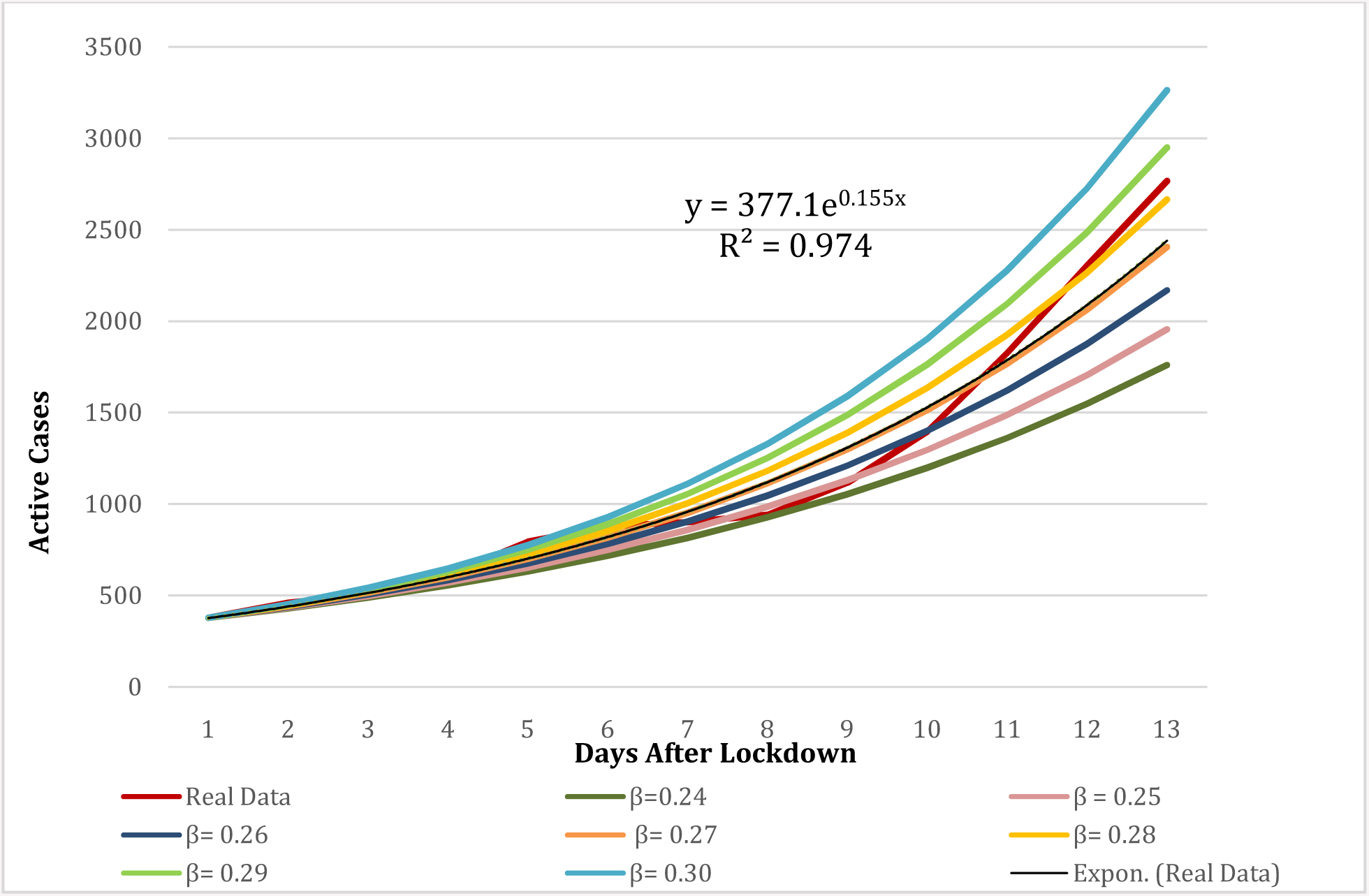
Figure 3. This graph compares real data with varying values of β, thus helps in estimating the range β lies in. Datasource:https://www.statista.com/statistics/1104054/india-coronavirus-covid-19-daily-confirmed-recovered-death-cases/.

Also visible in the graph, is the real data line shifting from 0.24 to 0.28 from day 8. Also, as R_t_ = β/**γ**, the value of the reproductive number is found to be varying between 2.4-2.9 in the Indian scenario until 4th April 2020. Considering the present trend of β, India can have nearly 7765 active cases by the end of the 21 days lockdown. Considering the range in which contact rate varies, number of active cases after 21 days lockdown period may vary from 5583 to 13785. (Figure3)

## DISCUSSION

After the disease set foot in India on 30th January, 2020, the government started taking precautionary measures to control the outbreak of the disease. These interventions included restricting the inflow of international passengers, self quarantine measures, directives on testing and management strategies in-country travel restrictions.

The median age of affected patients in our study sample of 413 was 36 years (IQR, 25-54) and higher in females (40 years) compared to males (35 years). In study cohorts of Wuhan, the median age of affected patients ranges from 49-56 years *(4, 23, 24)*. Thus, there are more people affected at a lower age in India when compared to China. This observation can be explained by the population distribution of India. In previous studies on COVID-19, it has been established that the risk increases with age and comorbidities *(13, 23, 24)*. However, according to the population demographics in India for 2020 *(25)*, the broad based nature of India’s population pyramid means there are more people in the younger age group and very few people in the equal to or above 80 years age group. This is reflected in the number of cases which are more in the younger age groups. On normalizing the percentage of patients in each age group with the corresponding percentage representation of the population, we observed that the highest number of male patients is in the 20-39 years age group category. Interestingly, according to this analysis, males in the 20-39 years age group are affected more than the 60-79 years age group males and equal to or above 80 years age group males. This is something that has not been reported until now and it has to be seen whether this changes as the number of cases in India grows. This is new to the literature of COVID-19 and needs to be studied further. Similarly finding the normalized ratio of affected patients by age group shows that for females, equal to or above 80 years age group is most affected.

As of 30 March, 2020, India has still not officially reported community spread *(26)*. So the cases now are people who were affected in foreign countries or their direct contacts. In 2018, Indian residents between 35 and 49 years of age took the most holidays outside the nation *(27)*. So, it is also possible that the number of young patients is more in India than other countries because they constitute the majority of outbound tourists. If community spread begins, this can get altered.

In our study, we found a case fatality rate of 3.87% which is lower than countries like Italy *(28)*. The explanation for a lower mortality rate due to COVID-19 infection could be the universal immunisation policy against BCG *(29)*. Case fatality rate was higher for males (4.38%) than females (2.88%). In China, the case fatality rate was found as 2.3%, 14.8% in the above 80 years population, and 8.0% in the 70-79 years population *(13)*. Our estimate gives a higher value which may be due a smaller sample size or can also be because mild cases of COVID 19 have so far been missed due to limited testing capacities in India. In our study, the mortality rates for 60-79 years age group (16.18%) and equal to or above 80 years age group (20%) may be an underestimation or overestimation of the actual mortality rate as the total number of deaths is still small and many are still hospitalized. The median age for recovered patients (36 years) is lower than the median ages for the deceased (65 years) implying younger patients have a higher chance of recovery. This is in accordance with findings from other countries *(13, 30)*. If we consider all the patients, males account for 66.34% of patients, implying more males are generally affected. 75.0% of the deceased were also males which show that males are more vulnerable agreeing with previous studies *(31, 32)*.

In the SIR model, the value of β being 0.258 (the range being between 0.24-0.29) and the trend for R_t_ being about 2.50, shows that the interventions which were put in place by the Indian government (assuming a lag period of 11 days) during the mid-March period were partially effective, preventing the scenario where Rt can reach even more than 4 *(33)*, and further data would tell how effective the lockdown is. The increase in β from day 8 is probably due to the identification of the Tablighi cluster and the cases it added to the Indian data *(34)*.

According to a study *(33)* in the early days of the epidemic, Wuhan city and Hubei province reported Rt between 1.85 and 4.46 which aligns with our study findings. All over China, the Rt varied from 1.23 to 5.77. South Korea which has high population density like India had a decreasing trend of Rt from 9.72 on 20 February to 1.50 on 7 March. This indicates that the interventions have been helpful in preventing the worst case scenario in India but is unable to prevent the spike in number of cases *(34,35)*.The situation can still be controlled if Rt can be brought down close to 1.This indicates the need for more effective strategies and ensuring optimum testing to avoid underestimation of danger.

The impact of the COVID-19 response (overall quarantine regulations, social distancing, and isolation of infections) in China in this context is encouraging for the many other countries *(36)* where the Rt gradually stabilized for most provinces ranging between 0.96 to 1.57 where there were more than 100 cases *(33)*, and India should try and replicate this when community transmission starts by strict enforcement of lockdown measures to get the Rt closer to 1,so that COVID would not overwhelm the Indian healthcare system. The Indian government will not be able to minimise both deaths from coronavirus disease 2019 (COVID-19) and the economic impact of viral spread. Keeping mortality as low as possible will be the highest priority for individuals; hence the Indian government must put in place measures to ameliorate the inevitable economic downturn *(37)*. However the Indian government’s timely decision to put a country wide lockdown into place when the number of cases was documented to be only 415,and effective contact tracing definitely helped to reduce the Rt and prevent an Italy/US like situation (however in the context of violations of the lockdown, more data awaited). Since at the end of the 21 days lockdown, India is likely to have significant number of active cases (5583 to 13785), it may enter the exponential growth phase and it would become very difficult to contain it. Hence, appropriate public health interventions like strict social distancing measures are necessary immediately to decrease the R_t_ in India.

The increase in β can be attributed to two possibilities: the Nizamuddin super cluster and change in the testing strategy. As on 4^th^ April,2020, 1023 COVID-19 positive cases across 17 states in India can be traced to the Tablighi Jamaat congregation in Nizamuddin *(38)*. Additionally, ICMR recently updated the testing strategy which might have increased the detection of the positive cases significantly *(39)*. For now the epidemic is being reported either in clusters or super-clusters and community transmission is still in its infancy. If the disease leaks into the rural areas or urban slums, it will provide a big challenge to the health care delivery system which is not yet ready to combat the epidemic. India might as well enter the exponential phase of growth of the epidemic curve if stringent measures are not in place. Hence, strict social distancing, optimum testing and acceptance and awareness in Indian community are vital elements to control the COVID-19 epidemic in India.

## LIMITATIONS

We have used crowd sourced data from covid19india.org due to lack of availability of official data. We could not include all the cases of COVID-19 till 28^th^ March due to unavailability of the demographic details of few patients.

## CONCLUSION

COVID-19 epidemic in India is affecting younger age groups more as compared to other countries.. Although the contact rate decreased initially after the Indian government’s stringent measures, the recent increase may be attributed to factors such as increase in testing or the occurrence of superclusters. Our mathematical model predicts that India will have adequate number of active cases after the 21 days lockdown. Hence, there is a need to develop appropriate public health interventions to control the COVID-19 epidemic in India.

## Data Availability

Data taken from publicly available crowdsourced and government databases

https://www.covid19india.org/

## ABBREVIATIONS

IQR: Interquartile Range
SIR: Susceptible-Infected-Removed

## COMPETING INTERESTS

None

## FUNDING

This research received no specific grant from any funding agency in the public, commercial or not-for-profit sectors

## ETHICS APPROVAL AND CONSENT TO PARTICIPATE

Mathematical modelling was done and available data in the public domain was used for analysis. Ethical approval was not required.

## ACKNOWLEDGEMENTS

We thank Hemant D Shewade of International Union against Tuberculosis and Lung Disease (The Union), Gowtham Balachandhiran of Panimalar Institute of Technology, Pidarithangal, Tamil Nadu, India, Krushna Prasad Pradhan of National Institute of Technology, Rourkela, Odisha, India and members of the Indian COVID Apex Research Team (i-CART) for their valuable feedback and sharing their expertise.

## BIOGRAPHICAL SKETCH

Mehak Arora, Archisman Mazumder, Vishwesh Bharadiya and Parul Berry are all medical students (MBBS) at All India Institute of Medical Sciences, New Delhi. Their research interests include epidemiology, public health and clinical investigations.

---

**Appendix Table:** Output of the SIR model run by varying β.

| Days after<br>lockdown | $\beta = 0.28$ | real data | $\beta = 0.29$ | $\beta = 0.30$ | $\beta = 0.27$ | $\beta = 0.26$ | $\beta = 0.25$ | $\beta = 0.24$ |
| --- | --- | --- | --- | --- | --- | --- | --- | --- |
| 0 | 378 | 378 | 378 | 378 | 378 | 378 | 378 | 378 |
| 1 | 444.8304 | 461 | 448.6104 | 452.3904 | 441.0504 | 437.2704 | 433.4904 | 429.7104 |
| 2 | 523.4764<br>147 | 507 | 532.4108<br>227 | 541.4208<br>307 | 514.6176<br>067 | 505.8343<br>987 | 497.1267<br>907 | 488.4947<br>827 |
| 3 | 616.0270<br>448 | 615 | 631.8651<br>644 | 647.9724<br>502 | 600.4558<br>235 | 585.1492<br>324 | 570.1050<br>036 | 555.3208<br>69 |
| 4 | 724.9406<br>264 | 794 | 749.8975<br>771 | 775.4934<br>284 | 700.6118<br>549 | 676.9006<br>321 | 653.7964<br>181 | 631.2887<br>639 |
| 5 | 853.1101<br>291 | 879 | 889.9784<br>445 | 928.1105<br>351 | 817.4739<br>123 | 783.0386<br>512 | 749.7737<br>323 | 717.6490<br>668 |
| 6 | 1003.94 | 902 | 1056.226<br>418 | 1110.762<br>688 | 953.8285<br>608 | 905.8191<br>117 | 859.8405<br>162 | 815.8234<br>591 |
| 7 | 1181.436<br>592 | 942 | 1253.529<br>513 | 1329.360<br>786 | 1112.927<br>165 | 1047.851<br>548 | 986.0651<br>04 | 927.4281<br>083 |
| 8 | 1390.314<br>581 | 1117 | 1487.688<br>826 | 1590.978<br>988 | 1298.563<br>416 | 1212.154<br>671 | 1130.819<br>461 | 1054.300<br>274 |
| 9 | 1636.122<br>199 | 1397 | 1765.589<br>098 | 1904.083<br>653 | 1515.163<br>794 | 1402.220<br>524 | 1296.823<br>758 | 1198.528<br>551 |
| 10 | 1925.388<br>604 | 1826 | 2095.401<br>142 | 2278.807<br>316 | 1767.893<br>114 | 1622.088<br>702 | 1487.197<br>486 | 1362.487<br>257 |
| 11 | 2265.797<br>309 | 2303 | 2486.822<br>075 | 2727.276<br>596 | 2062.777<br>686 | 1876.432<br>21 | 1705.518<br>077 | 1548.875<br>513 |
| 12 | 2666.390<br>274 | 2767 | 2951.360<br>439 | 3264.004<br>63 | 2406.849<br>004 | 2170.656<br>781 | 1955.888<br>13 | 1760.761<br>684 |
| 13 | 3137.808<br>074 |  | 3502.674<br>569 | 3906.360<br>741 | 2808.311<br>418 | 2511.015<br>764 | 2243.012<br>508 | 2001.633<br>882 |
| 14 | 3692.572<br>542 |  | 4156.974<br>179 | 4675.132<br>535 | 3276.737<br>762 | 2904.743<br>036 | 2572.286<br>744 | 2275.457<br>397 |
| 15 | 4345.419<br>367 |  | 4933.496<br>955 | 5595.198<br>617 | 3823.297<br>621 | 3360.206<br>744 | 2949.898<br>438 | 2586.739<br>969 |
| 16 | 5113.689<br>511 |  | 5855.074<br>186 | 6696.333<br>705 | 4461.023<br>664 | 3887.087<br>161 | 3382.943<br>529 | 2940.605<br>997 |
| 17 | 6017.789<br>817 |  | 6948.802<br>045 | 8014.172<br>178 | 5205.122<br>411 | 4496.582<br>428 | 3879.559<br>639 | 3342.880<br>897 |
| 18 | 7081.735 |  | 8246.838 | 9591.361 | 6073.336 | 5201.646 | 4449.078 | 3800.187 |
|  | 056 |  | 266 | 263 | 83 | 553 | 994 | 004 |
| 19 | 8333.785 |  | 9787.347 | 11478.94 | 7086.369 | 6017.264 | 5102.203 | 4320.052 |
|  | 814 |  | 655 | 116 | 413 | 732 | 79 | 586 |
| 20 | 9807.199 |  | 11615.62 | 13737.99 | 8268.375 | 6960.771 | 5851.207 | 4911.035 |
|  | 146 |  | 42 | 678 | 831 | 842 | 307 | 78 |
| 21 | 11541.11 |  | 13785.42 | 16441.63 | 9647.540 | 8052.220 | 6710.164 | 5582.865 |
|  | 196 |  | 28 | 455 | 919 | 867 | 539 | 474 |

## Notes

### Competing Interest Statement

The authors have declared no competing interest.

### Funding Statement

No funding recieved

## REFERENCES

1. Zhu N, Zhang D, Wang W, Li X, Yang B, Song J, et al. A Novel Coronavirus from Patients with Pneumonia in China, 2019. N Engl J Med. 2020 Feb 20;382(8):727–33.

2. Tang X, Wu C, Li X, Song Y, Yao X, Wu X, et al. On the origin and continuing evolution of SARS-CoV-2. Natl Sci Rev.[cited 2020 Apr 2]; Available from: https://f1000.com/prime/737471750.

3. WHO Director-General’s opening remarks at the media briefing on COVID-19 - 11 March 2020 [Internet]. [cited 2020 Apr 2]. Available from: https://www.who.int/dg/speeches/detail/who-director-general-s-opening-remarks-at-the-media-briefing-on-covid-19-11-march-2020.

4. Guan W, Ni Z, Hu Y, Liang W, Ou C, He J, et al. Clinical Characteristics of Coronavirus Disease 2019 in China. N Engl J Med [Internet]. 2020 Feb 28 [cited 2020 Apr 2]; Available from: http://www.nejm.org/doi/10.1056/NEJMoa2002032.

5. Modes of transmission of virus causing COVID-19: implications for IPC precaution recommendations [Internet]. [cited 2020 Apr 2]. Available from: https://www.who.int/news-room/commentaries/detail/modes-of-transmission-of-virus-causing-covid-19-implications-for-ipc-precaution-recommendations.

6. van Doremalen N, Bushmaker T, Morris DH, Holbrook MG, Gamble A, Williamson BN, et al. Aerosol and Surface Stability of SARS-CoV-2 as Compared with SARS-CoV-1. N Engl J Med. 2020 Mar 17.

7. Lu S, Lin J, Zhang Z, Xiao L, Jiang Z, Chen J, et al. Alert for non-respiratory symptoms of Coronavirus Disease 2019 (COVID-19) patients in epidemic period: A case report of familial cluster with three asymptomatic COVID-19 patients. J Med Virol [Internet]. 2020 Mar 19 [cited 2020 Mar 29]; Available from: http://doi.wiley.com/10.1002/jmv.25776.

8. Rothe C, Schunk M, Sothmann P, Bretzel G, Froeschl G, Wallrauch C, et al. Transmission of 2019-nCoV Infection from an Asymptomatic Contact in Germany. N Engl J Med. 2020 05;382(10):970–1.

9. Qian G, Yang N, Ma AHY, Wang L, Li G, Chen X, et al. A COVID-19 Transmission within a family cluster by presymptomatic infectors in China. Clin Infect Dis [Internet]. 2020 Mar 23 [cited 2020 Mar 29]; Available from: https://academic.oup.com/cid/advance-article/doi/10.1093/cid/ciaa316/5810900.

10. Chan JF-W, Yuan S, Kok K-H, To KK-W, Chu H, Yang J, et al. A familial cluster of pneumonia associated with the 2019 novel coronavirus indicating person-to-person transmission: a study of a family cluster. The Lancet. 2020 Feb 15;395(10223):514–23.

11. Li Q, Guan X, Wu P, Wang X, Zhou L, Tong Y, et al. Early Transmission Dynamics in Wuhan, China, of Novel Coronavirus–Infected Pneumonia. N Engl J Med. 2020 Mar 26;382(13):1199–207.

12. Young BE, Ong SWX, Kalimuddin S, Low JG, Tan SY, Loh J, et al. Epidemiologic Features and Clinical Course of Patients Infected With SARS-CoV-2 in Singapore. JAMA. 2020 1. Mar 3.

13. Wu Z, McGoogan JM. Characteristics of and Important Lessons From the Coronavirus Disease 2019 (COVID-19) Outbreak in China: Summary of a Report of 72 314 Cases From the Chinese Center for Disease Control and Prevention. JAMA [Internet]. 2020 Feb 24 [cited 2020 Apr 2]; Available from: https://jamanetwork.com/journals/jama/fullarticle/2762130.

14. Adaptive COVID-19 Treatment Trial (ACTT) - Full Text View - ClinicalTrials.gov [Internet]. [cited 2020 Apr 2]. Available from: https://clinicaltrials.gov/ct2/show/NCT04280705.

15. COVID 100casesIndia [Internet]. StoryMapJS. [cited 2020 Apr 2]. Available from: https://uploads.knightlab.com/storymapjs/62428320e6fd66f08eb61ab6b26cd60c/covid-100casesindia/draft.html.

16. Ministry of health and Family Welfare, Government of India [Internet]. [cited 2020 Apr 1]. Available from: https://www.mohfw.gov.in/.

17. COVID-19 Tracker | India [Internet]. [cited 2020 Apr 2]. Available from: https://www.covid19india.org.

18. Global surveillance for COVID-19 caused by human infection with COVID-19 virus. Interim Guidance. [Internet]. WHO; 2020 [cited 2020 Apr 2]. Available from: https://www.who.int/docs/default-source/coronaviruse/global-surveillance-for-covid-v-19-final200321-rev.pdf.

19. Kermack WO, McKendrick AG, Walker GT. A contribution to the mathematical theory of epidemics. Proc R Soc Lond Ser Contain Pap Math Phys Character. 1927 Aug 1;115(772):700–21.

20. Huppert A, Katriel G. Mathematical modelling and prediction in infectious disease epidemiology. Clin Microbiol Infect. 2013 Nov;19(11):999–1005.

21. COVID-19 India [Internet]. [cited 2020 Apr 2]. Available from: http://covidindiaupdates.in/.

22. India - daily cumulative COVID-19 by type 2020 [Internet]. Statista. [cited 2020 Apr 2]. Available from: https://www.statista.com/statistics/1104054/india-coronavirus-covid-19-daily-confirmed-recovered-death-cases/.

23. Huang C, Wang Y, Li X, Ren L, Zhao J, Hu Y, et al. Clinical features of patients infected with 2019 novel coronavirus in Wuhan, China. The Lancet. 2020 Feb;395(10223):497–506.

24. Chen N, Zhou M, Dong X, Qu J, Gong F, Han Y, et al. Epidemiological and clinical characteristics of 99 cases of 2019 novel coronavirus pneumonia in Wuhan, China: a descriptive study. Lancet Lond Engl. 2020 15;395(10223):507–13.

25. Population Pyramids of the World from 1950 to 2100 [Internet]. PopulationPyramid.net. [cited 2020 Apr 2]. Available from: https://www.populationpyramid.net/india/2020/.

26. India coronavirus latest update, March 30: Centre says ‘no community transmission’ yet [Internet]. The Indian Express. 2020 [cited 2020 Apr 2]. Available from: https://indianexpress.com/article/coronavirus/india-coronavirus-lockdown-updates-march-30-6338319/.

27. India Tourism Source (Domestic & Outbound) Market Insights 2019 - ResearchAndMarkets.com [Internet]. 2019 [cited 2020 Apr 2]. Available from: https://www.businesswire.com/news/home/20191015005520/en/India-Tourism-Source-Domestic-Outbound-Market-Insights.

27. Dandona L, Dandona R, Kumar GA, Shukla DK, Paul VK, Balakrishnan K, et al. Nations within a nation: variations in epidemiological transition across the states of India, 1990– 2016 in the Global Burden of Disease Study. The Lancet. 2017 Dec 2;390(10111):2437–60.

28. Livingston E, Bucher K. Coronavirus Disease 2019 (COVID-19) in Italy. JAMA [Internet]. 2020 Mar 17 [cited 2020 Apr 6]; Available from: https://jamanetwork.com/journals/jama/fullarticle/276340.

29. Miller A, Reandelar MJ, Fasciglione K, Roumenova V, Li Y, Otazu GH. Correlation between universal BCG vaccination policy and reduced morbidity and mortality for COVID-19: an epidemiological study. medRxiv. 2020 Mar 28;2020.03.24.20042937.

30. Onder G, Rezza G, Brusaferro S. Case-Fatality Rate and Characteristics of Patients Dying in Relation to COVID-19 in Italy. JAMA. 2020 Mar 23.

31. Novel Coronavirus Pneumonia Emergency Response Epidemiology Team. [The epidemiological characteristics of an outbreak of 2019 novel coronavirus diseases (COVID-19) in China]. Zhonghua Liu Xing Bing Xue Za Zhi Zhonghua Liuxingbingxue Zazhi. 2020 Feb 17;41(2):145–51.

32. World Health Organization. Report of the WHO-China Joint Mission on Coronavirus Disease 2019 (COVID-19) [Internet]. World Health Organization; 2020 [cited 2020 Apr 2]. Available from: https://www.who.int/docs/default-source/coronaviruse/who-china-joint-mission-on-covid-19-final-report.pdf.

33. Huang Y, Yang L, Dai H, Tian F, Chen K. Epidemic situation and forecasting of COVID-19 in and outside China. Bull World Health Organ. (E-pub: 16 March 2020).

34. Philip SA. How Tablighi Jamaat emerged as the “largest known” Covid-19 source in South Asia [Internet]. ThePrint. 2020 [cited 2020 Apr 2]. Available from: 27. https://theprint.in/india/how-tablighi-jamaat-unknowingly-emerges-as-largest-known-covid-19-source-in-south-asia/391918/.

35. Huge crowds as lockdown sparks mass migration [Internet]. BBC News. [cited 2020 Apr 2]. Available from: https://www.bbc.com/news/av/world-asia-india-52093574/coronavirus-huge-crowds-as-india-lockdown-sparks-mass-migration.

36. Rm A, H H, D K, Td H. How will country-based mitigation measures influence the course of the COVID-19 epidemic? The Lancet [Internet]. 2020 Mar 6 [cited 2020 Apr 3]; Available from: https://www.bdi.ox.ac.uk/publications/1093310.

37. Coronavirus to impact India’s economic growth “severely”: D&B. The Economic Times [Internet]. 2020 Mar 27; Available from: https://economictimes.indiatimes.com/news/economy/indicators/coronavirus-to-impact-indias-economic-growth-severely-db/articleshow/74825429.cms?from=mdr.

38. 1,023 COVID-19 positive cases with links to Tablighi Jamaat reported from 17 states: Health ministry [Internet]. Deccan Herald. 2020 [cited 2020 Apr 7]. Available from: https://www.deccanherald.com/national/north-and-central/1023-covid-19-positive-cases-with-links-to-tablighi-jamaat-reported-from-17-states-health-ministry-821321.html.

39. Ministry of health and Family Welfare, Government of India [Internet]. [cited 2020 Apr 1]. Available from: https://www.mohfw.gov.in/.

